# Longitudinal monitoring of SARS-CoV-2 neutralizing antibody titers and its impact on employee personal wellness decisions

**DOI:** 10.1101/2022.03.01.22271202

**Authors:** Rianna Vandergaast, Timothy Carey, Lukkana Suksanpaisan, Chase Lathrum, Riya Narjari, Michelle Haselton, Luke Schnebeck, Aroshi Wijesekara, Andrew Duncan, Luke Russell, Shruthi Naik, Kah-Whye Peng, Patrycja Lech, Stephen J. Russell

## Abstract

Virus neutralizing antibody (vnAb) titers are the strongest laboratory correlate of protection from SARS-CoV-2. Providing individuals with real-time measures of their vnAb titers is predicted to improve their ability to make personal wellness decisions. Yet, widespread commercial testing of SARS-CoV-2 vnAbs does not currently occur. Here, we examined whether knowing their vnAb titer impacted wellness decision-making among individuals. To this end, starting on January 1, 2021, we offered all employees from two companies free IMMUNO-COV™ testing and conducted a survey to assess their behaviors and decisions regarding booster vaccination. IMMUNO-COV is a clinically validated, surrogate virus assay that quantitates serum titers of SARS-CoV-2 vnAbs. To help participants gauge their level of protection based on their vnAb titer, we calibrated IMMUNO-COV titers to the World Health Organization (WHO) International Standard (IU/mL), making them comparable to published reports of correlates of protection, and we fit historical IMMUNO-COV vnAb titer values into predictive models of immune protection from COVID-19. As expected, data for the 56 program participants showed variability in vnAb titers post vaccination, rates vnAb decay, and fold-increases in vnAb titers after booster vaccination. Based on the participant survey, the majority (66%) of participants indicated that knowing their vnAb titer impacted their social behaviors and/or their decision on the timing of a booster vaccination. Several participants indicated that knowing their vnAb titer contributed to their peace of mind regarding their high level of protection from COVID-19. Together, these data demonstrate that regular determination of SARS-CoV-2 neutralizing antibody titers can significantly impact decisions regarding social interactions and timing of booster vaccinations.

## INTRODUCTION

Over the course of the COVID-19 pandemic, people have continually made behavioral and personal wellness decisions in the context of frequently changing infection and hospitalization rates, available scientific information, and regulatory recommendations or mandates. Central to these decisions is an understanding of how a person’s actions impact their risk of becoming infected with SARS-CoV-2 or spreading SARS-CoV-2 to others. For the most vulnerable populations – the elderly, immune compromised, and those with other comorbid conditions – and their close contacts, accurately assessing their risk of COVID-19 has been especially important and challenging. Beyond an understanding of risk factors, few tools have been effectively deployed to inform individuals of their personal risk of contracting SARS-CoV-2. Recommendations and mandates, including lockdowns, quarantining, social distancing, masking, and vaccination, have been used by governments and health agencies to help protect citizens, primarily the most vulnerable, from COVID-19. Yet, people have limited options for making personalized risk assessments based on knowledge of their level of protection from SARS-CoV-2 infection afforded by vaccination or previous COVID-19 disease.

In 2021, several key studies indicated that SARS-CoV-2 virus neutralizing antibody (vnAb) titers provide the best correlate of protection from future infection, correlating strongly with vaccine efficacy for both mRNA and adenovirus vaccines and providing the best predictor of individual risk of infection^1–4^. Breakthrough infections, which occur most commonly with SARS-CoV-2 variants, coincide with low vnAb titers in individuals and with overall waning immunity in some vaccinated populations^5–7^. Therefore, when individuals know their vnAb titer, they can more accurately predict their level of protection from future SARS-CoV-2 infection. When considered in combination with their medical history, this information can help them more precisely evaluate the risks and benefits of their behaviors and medical decisions.

Numerous serological assays for detecting SARS-CoV-2 antibodies have been developed. However, most are qualitative, do not look specifically for neutralizing antibodies, or are for research use only. Thus, patient access to vnAb testing is limited. IMMUNO-COV was developed as a collaboration between Vyriad, Inc., Regeneron Pharmaceuticals, and Imanis Life Sciences^8^. It is a clinically validated laboratory developed test (LDT) that measures SARS-CoV-2 vnAb titers in blood (serum or plasma) samples. Initially launched in April 2020, IMMUNO-COV was the first commercially scalable test of its kind, available to patients through physician order for accurate measure of an individual’s vnAb titer.

In early 2021, following the rollout of vaccination programs and the launch of IMMUNO-COV v2.0^9^, free testing was offered to all employees from two companies. This free testing provided employees the opportunity to measure their level of protection from COVID-19 in real-time following vaccination or recovery from infection, allowing them to personally assess the risks of their behaviors during the COVID-19 pandemic. During the period of this study, the dominant SARS-CoV-2 variant in the U.S.A. evolved from Wuhan (ancestral) to Delta (B.1.617.2). Here, we report on our retrospective study of individuals involved in the free IMMUNO-COV testing program. Of the employees that participated in the program from January 1 through December 4, 2021, we had data from 56 evaluable participants encompassing the time from first vaccination series through booster vaccination. To correlate IMMUNO-COV titers with published reports and help participants more easily estimate their level of protection from their test results, we calibrated the IMMUNO-COV assay to the World Health Organization (WHO) International Standard Unit (IU/mL)^10^. We also applied published models correlating vnAb titers and protection levels^1,2^ to IMMUNO-COV historical data to generate models of protection for SARS-CoV-2 Wuhan and Delta. Using the data collected from participants, we examined the rate of vnAb decline, the correlation between vnAb levels and age, and the fold change in titers following booster vaccination. Finally, we conducted a voluntary questionnaire survey to evaluate the impact that knowing vnAb titer had on participant life choices during the COVID-19 pandemic.

## MATERIALS AND METHODS

### Study Design and Population

The study protocol, informed consent forms to collect blood, and participant survey were reviewed and approved by Western Institutional Review Board. Written informed consent was obtained from all study participants. We are reporting data collected from January 1 through December 4, 2021. IMMUNO-COV was offered free to all employees from two selected companies. Participants provided blood samples for IMMUNO-COV testing at intervals of their choosing before and after vaccination. Participants were notified of their personal test results 1-2 weeks after taking the test. Approximately 90% of employees participated in the study, all of whom received at least two doses of BNT162b2 vaccine (Pfizer-BioNTech) or mRNA-1273 vaccine (Moderna) or one dose of Ad26.CoV2.S (Janssen) during the study course. The study was performed retrospectively to characterize peak vnAb titers post vaccination and booster vaccination and examine the kinetics of vnAb decay.

### Data Collection

Participants completed a questionnaire that provided de-identified demographic information including age and gender, and information relevant to their COVID-19 immunity including date of first, second, and booster vaccination and the vaccine(s) that they received (see in part Table 1). Participants also informed us if they had COVID-19 prior to vaccination or a breakthrough infection following vaccination and provided the date of their COVID-19 test which detected SARS-CoV-2 by RT-PCR assay. Near the end of the study, participants were emailed a request to complete a survey, which was securely completed through DocuSign.

**Table 1:** Study Population and Demographics

| **Group** | **Number** |
| --- | --- |
| **Total number of participants** | 56 |
| **Gender** |  |
| Male | 26 (46%) |
| Female | 30 (54%) |
| **Mean age in years (range)** |  |
| Male | 42 (22-70) |
| Female | 40 (22-64) |
| **Age group (years)** |  |
| 20-30 | 14 (25%) |
| 31-40 | 18 (32%) |
| 41-50 | 6 (11%) |
| 51-60 | 15 (27%) |
| 61+ | 3 (5%) |
| **COVID-19 positive before vaccination** | 4 (7%) |
| **COVID-19 positive after vaccination** | 2 (4%) |
| **Vaccine regimen (number)** |  |
| Pfizer (X3) | 32 (57%) |
| Pfizer (X2) | 14 (25%) |
| Moderna (X2) | 4 (7%) |
| Moderna (X3) | 3 (5%) |
| Pfizer (X2), Janssen (X1) | 2 (4%) |
| Janssen (X1) | 1 (2%) |

### Data Analysis and Statistics

Data was graphed and analyzed using Prism v9.3.1 (GraphPad Prism, San Diego, CA). Correlation analysis was performed with log-transformed data using Pearson’s correlation and simple linear regression with 95% confidence interval. Because the frequency of sample collection/testing varied between participants, with sample numbers ranging from one to ten per participant, we calculated mean average titers per two-month intervals and performed comparisons between each two-month interval. If a participant had two or more collection days in a time interval, only the highest vnAb titer was used. Collection days following booster vaccine or a breakthrough infection by SARS-CoV-2 were omitted for analyses of the kinetics of vnAb decay.

### Study Limitations

This was a retrospective study with a sample size too small for statistical inter-group comparisons. Sample collection was not stratified; blood samples for testing were not collected at the same intervals relative to vaccination for all participants and the number of data collection points varied between individuals. Only 16/56 (29%) participants had titers measured within 8-30 days post second vaccination, meaning the peak titers recorded for many participants likely underestimate their true peak titer. All correlates for protection are based on vnAb levels, which provide the best correlate of protection from future infection^1,2^. However, cellular immunity also plays a role in protecting against SARS-CoV-2, and cellular immune responses were not examined as part of this study.

### IMMUNO-COV Assay

Quantitation of vnAb titers was performed using IMMUNO-COV v2.0 (referred to throughout this manuscript as IMMUNO-COV)^9^. The clinically validated test detects the presence of specific neutralizing antibodies capable of inhibiting a vesicular stomatitis virus (VSV)-based surrogate virus, VSV-SARS2-Fluc. Infection by VSV-SARS2-Fluc results in the expression of firefly luciferase within infected ACE2-expressing cells and total luminescence corresponds to the extent of infection by VSV-SARS2-Fluc. The relative neutralizing capacity present in an unknown sample is inversely proportional to the luminescence and is derived by reference to a standard curve. For most of the study period, vnAb titers were reported as an IMMUNO-COV-specific virus neutralizing titer (VNT) that correlates closely to results produced by a traditional plaque reduction neutralization test (PRNT). Near the end of the study, IMMUNO-COV titers were standardized to the World Health Organization (WHO) International Standard unit (IU/mL) and data presented here were retrospectively converted to IU/mL, making them comparable to titers obtained using other standardized neutralization assays.

### IMMUNO-COV Delta (B.1.617.2) Assay

VSV-SARS2(Delta)-Fluc virus was generated by replacing the SΔ19CT spike glycoprotein (Wuhan) from VSV-SARS2-Fluc^9^ with SΔ19CT spike glycoprotein Delta (mutations compared to Wuhan: T19R, E156-, F157-, R158G, L452R, T478K, P681R, D950N). Plasmid was sequence verified and used for infectious virus rescue on BHK-21 cells as previously described^11^. The rescued virus was amplified in Vero-ACE2 cells^9^ to generate large-scale stocks for use in the assay. For vnAb assay, test serum samples at increasing dilutions were incubated with VSV-SARS2(Delta)-Fluc (800 pfu/100 µL) for 30 min at room temperature, before being overlaid onto monolayers of Vero-ACE2 cells plated 16-24 h before use. After an additional 28 h, d-luciferin substrate was added to wells and bioluminescence was measured using a Tecan M Plex or Tecan Lume instrument (100 ms integration, 100 ms settle time per well). Pooled seronegative serum control was included on every assay plate and the relative luciferase signal (response) in wells for each sample dilution were normalized to the luciferase signal from the negative control wells. Percent response values were plotted against dilution factors and regression analyses were performed. A 50% inhibitory concentration (IC50) titer was determined from the fitted curves for each sample. For experiments comparing vnAb titers against SARS-CoV-2 Wuhan and Delta, the original IMMUNO-COV v2.0 assay was run in a similar IC50 format, and IC50 titers were determined rather than typical IMMUNO-COV VNT or IU/mL titers.

### Standardization of IMMUNO-COV to the WHO IU/mL

For standardization, the First WHO International Reference Panel for anti-SARS-CoV-2 Immunoglobulin (NIBSC code 20/268) and the SARS-CoV-2 National Standard (Frederick National Labs, calibrated separately to the WHO International Standard) were used as sample basis. The WHO Reference Panel contains five pooled plasma samples that range from negative to high positive vnAb titer, while the Frederick National Labs (FNL) Standard consists of a pooled plasma sample with a titer between that of the two highest WHO Reference Panel samples. Reference samples were assayed in triplicate over the course of four nonconsecutive days by four analysts resulting in a total of 15 unique runs using three distinct vials of each provided reference sample. Positive reference samples were assayed in eight-point, two-fold dilution series starting at 1:80 (WHO Panel) or 1:40 (FNL). The negative reference sample was tested at 1:80 only, which is the starting dilution of the standard IMMUNO-COV assay. Each assay plate also included the IMMUNO-COV standard curve, consisting of stepped concentrations of neutralizing monoclonal antibody mAb10914, which was used to calculate the IMMUNO-COV VNT of each reference sample. A second neutralizing monoclonal antibody, mAb10922, was also used for comparisons. Percent luciferase responses normalized to a negative control of pooled seronegative serum (included on every assay plate) were determined for each dilution, and dilutions that resulted in a luciferase signals that fell within the dynamic range of the assay were used to perform parallelism analyses of the reference samples and monoclonal antibody controls. Regression analyses and curve fitting (4-parameter logistic) were performed using Prism. From these analyses, the Hill slope was confirmed to be the same (F value = 1.433, *p* value = 0.2627). The neutralizing potency in IU/mL for each Reference Standard was provided (Table 2) and plotted against the corresponding mean geometric VNT titer results (from dilutions within the linear range) from the IMMUNO-COV assay for regression analyses. Several regression models were considered, but for the ease of performing data conversion, a linear regression with no intercept model was chosen (r^2^ = 0.9080). Based on this analysis, the correction factor for conversion of VNT values to WHO IU/mL values was 0.6954.

**Table 2:** Comparison of IMMUNO-COV and cPass titers (results) to nominal concentrations of Reference Samples

| **WHO Reference Sample** | **WHO Reference Value (IU/mL)** | **IMMUNO-COV** |  | **cPass** |  |
| --- | --- | --- | --- | --- | --- |
|  |  | **Result** | **Titer (IU/mL)** | **Result** | **Inhibition Factor\*** |
| Negative (20/142) | Negative | Negative | < LOD | Negative | -4.5 |
| Low (20/140) | 44 | Borderline | 32† | Negative | 16.3 |
| LowS (20/144) | 95 | Positive | 80 | Positive | 31.4 |
| Mid (20/148) | 210 | Positive | 280 | Positive | 70.9 |
| FNL | 813 | Positive | 861 | ND | ND |
| High (20/150) | 1473 | Positive | 1361 | Positive | 85.4 |

†Value from runs in which sample tested positive.
\*According to cPass kit instructions, samples with an inhibition factor above 30 are considered positive.
ND = not done

### Generation of Models of Protection

The predictive models utilized in this study were based on data and algorithms provided in the primary source publications and were obtained from the GitHub repository associated with each primary source.^1,2^ The source models were generated to evaluate protection based on the mean neutralization level as a fold-change of convalescent neutralization levels. Therefore, the adaptation of these models to evaluate protection of SARS-CoV-2, both ancestral (wildtype) and Delta based on IMMUNO-COV titers, required obtaining the mean convalescent titer from ancestral virus. Data collected early in the launch of the assay, prior to vaccines becoming readily available, was analyzed and although the specific details as to the timing of convalescence was not known in all cases, a mean convalescent titer was calculated from the geometric mean of 264 convalescent samples. The geometric mean of convalescent titers measured by the IMMUNO-COV, which was applied to the predictive models was 325.6 IU/mL (95% CI: 289.7 to 366 IU/mL). Transforming the fold-change of mean convalescent to this measured value from the IMMUNO-COV datasets yielded the models of protection established in this report and were also applied to Delta protection model according to similar assumptions set in the source models of protection. Prior reporting for models of protection using WHO IU/mL values is still limited at the time of publication, but Khourey *et al*. reported that 50% protection against symptomatic disease from SARS-CoV-2 Wuhan infection corresponds to 54 IU/mL^1^. Using the IMMUNO-COV predictive models, a similar titer result of 67 IU/mL was calculated to yield 50% protection against SARS-CoV-2 Wuhan infection.

## RESULTS

### Study Population and Demographics

A total of 214 serum samples were collected from 56 participants post primary vaccination against SARS-CoV-2 (Table 1). Participants provided blood samples at intervals of their choosing and did not adhere to a predetermined sample collection schedule. Longitudinal participation varied between individuals, with participants electing to receive IMMUNO-COV testing between 1 to 10 times during the study period (January 1, 2021, through December 4, 2021). The gender distribution of participants was near equal with 26/56 (46%) males (mean age, 42 years) and 30/56 (54%) females (mean age, 40 years). The age of the study population ranged from 22 to 70 years of age, with 14/56 (25%) participants aged 20-30 years, 18/56 (32%) aged 31-40 years, 6/56 (11%) aged 41-50 years, 15/56 (27%) aged 51-60 years, and 3/56 (5%) aged 61 years or older.

All participants were fully vaccinated, receiving at least two doses of BNT162b2 (Pfizer-BioNTech) or mRNA-1273 (Moderna) or one dose of Ad26.CoV2.S (Janssen) vaccines (Table 1). During the study period, 37/56 (66%) participants received a booster vaccine, of which 24/56 (43%) had pre- and post-booster vaccine samples collected. Booster doses were BNT162b2 (32/37), mRNA-1273 (3/37), or Ad26.CoV.S (2/37), and were received as early as five months post primary vaccination. Four participants (7%) tested positive for COVID-19 prior to the start of the study and receipt of their primary vaccine, while two participants (4%) had break-through infections following vaccination.

### Calibration of IMMUNO-COV to WHO International Standard

To readily compare IMMUNO-COV vnAb titers from this study with other published reports of correlates of protection, we calibrated IMMUNO-COV titers to the World Health Organization International Standard unit (IU/mL)^10^. To this end, we ran samples from the First WHO International Reference Panel for anti-SARS-CoV-2 Immunoglobulin (NIBSC code 20/268) and the SARS-CoV-2 National Standard (FNL) in multiple analytical runs on the IMMUNO-COV assay. Together, these reference samples include one negative sample and five positive samples ranging from low to moderately high titer (Table 2). Samples were assayed in triplicate over the course of four nonconsecutive days by four different analysts, resulting in a total of 15 unique runs. Monoclonal antibody calibrators mAb10914 and mAb10922^9^ were also included in comparisons. Positive reference samples were assayed in two-fold dilution series and percent luciferase responses (normalized to a negative control of pooled seronegative serum) were determined for each dilution. Percent response values for dilutions that fell within the dynamic range of the assay were plotted for regression analyses (Figure 1A), which confirmed the samples diluted in a parallel manner. Geometric mean VNT titers were determined for each reference sample based on the dilutions that were in the linear range and were plotted against the corresponding established potency values (IU/mL). Several regression models were considered, but for the ease of performing data conversion, a linear regression with no intercept model was chosen (r^2^ = 0.9080). Based on this analysis, the correction factor for conversion of VNT values to WHO IU/mL values is 0.6954.

**Figure 1.**
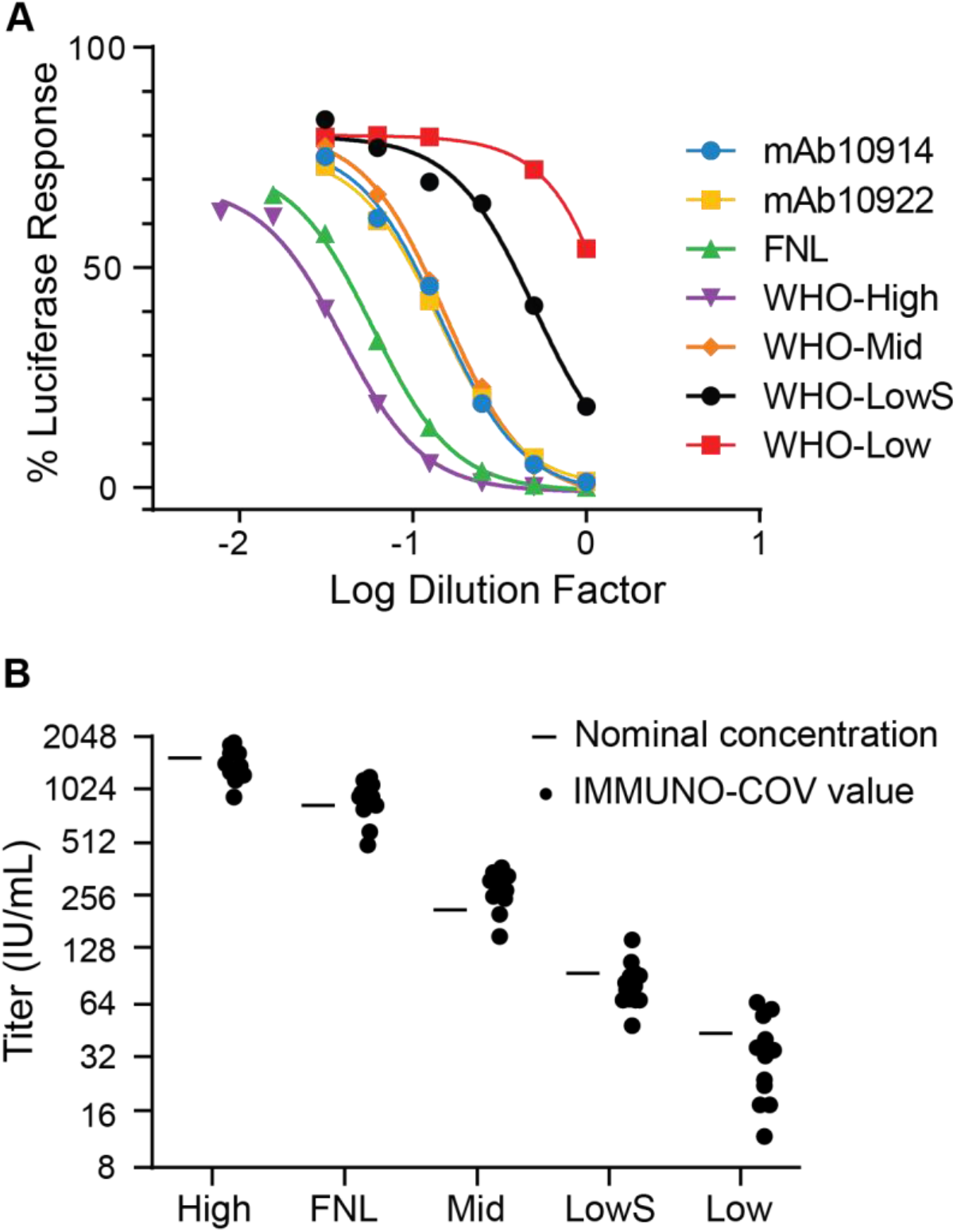
Standardization of IMMUNO-COV to WHO IU/mL. A) Regression fit curves. WHO Reference Samples (WHO-High, WHO-Mid, WHO-LowS, and WHO-Low), FNL Standard, and IMMUNO-COV monoclonal antibody calibrators mAb10914 and mAb10922 were assayed in four days of combined analytical runs (n = 15) using the IMMUNO-COV v2.0 assay. Regression curves were generated based on normalized luciferase signals of sample/control dilutions that fell within the linear range of the assay. Curve fits show that each calibrator material dilutes in a parallel manner (Hill slope was confirmed to be the same (F value = 1.433, p value = 0.2627)), allowing for a single-factor conversion for potency. B) Performance of calibrated IMMUNO-COV assay. Data from the individual runs described in Panel A, were converted to WHO Standard units (IU/mL) using the determined conversion factor of 0.6954. Dots represent individual titers of reference samples determined from each analytical run. Dashes represent the nominal neutralizing potency values provided from NIBSC or FNL.

To further assess the accuracy of the calibration and assay performance, the 0.6954 conversion factor was used to convert all VNT titer values from the 15 analytical runs to WHO IU/mL units, which were then plotted relative to the nominal concentrations of each sample (Figure 1B). From this analysis, IMMUNO-COV analytical runs demonstrated consistent performance with respect to the expected nominal concentrations, indicating appropriate calibration of IMMUNO-COV titer to the WHO International Standard. Unlike IMMUNO-COV, the cPass SARS-CoV-2 surrogate virus neutralization assay (GenScript)^12^ had lower sensitivity and poor quantitation when used to measure the Reference Samples (Table 2). This data further emphasizes the accuracy of the functional IMMUNO-COV assay relative to a neutralization assay that relies on binding alone to measure vnAb titers.

### Modeling protection of SARS-CoV-2 Wuhan and Delta

To help participants interpret vnAb titer results, we applied a predictive model of immune protection from COVID-19 to IMMUNO-COV historical data. Integrating published data on *in vitro* neuralization titers and clinical protection, the model described by Khourey *et al*. and Cormer *et al*.^1,2^ estimates 50% protection against SARS-CoV-2 Wuhan infection or severe disease to be 20% or 3%, respectively, of the mean titer from convalescent samples. All study participants received vaccines directed against the SARS-CoV-2 Wuhan spike glycoprotein. For SARS-CoV-2 Delta (B.1.617.2), which became the dominate strain in the U.S.A. approximately halfway through our study and contains several mutations in the spike region, the model applies a correction factor of 3.9 that represents the mean drop in neutralization titer between SARS-CoV-2 Wuhan and Delta^2^.

We also directly examined the relative *in vitro* potency of vaccine-induced vnAbs for SARS-CoV-2 Wuhan and Delta using IMMUNO-COV. To this end, we developed a modified IMMUNO-COV assay that uses VSV-SARS2(Delta)-Fluc to specifically measure vnAb titers against the Delta variant. IMMUNO-COV Delta assays were performed by incubating increasing dilutions of test samples with VSV-SARS2(Delta)-Fluc, encoding SARS-CoV-2 spike glycoprotein with the Delta variant sequence, overlaying the virus/serum mixes onto ACE2-overespressing cells, and after 28 h reading luciferase activity in the wells. The luciferase signals at each dilution relative to negative control serum were used to determine a 50% inhibitory concentration (IC50). To facilitate comparisons with Wuhan neutralization, for this assessment, the IMMUNO-COV assay was also run using dilution series to calculate IC50 titers. Three seropositive samples from previously vaccinated, non-convalescent participants were tested side-by-side in the Wuhan and Delta assays. For each sample, the luciferase response curves for Wuhan were clearly shifted toward higher dilutions (higher IC50 values) compared to luciferase response curves for Delta (Figure 2A). The degree of this shift ranged from 2.2 to 3.4-fold. Overall, the consensus curves demonstrated a shift of 2.76-fold (Figure 2B), indicating that vaccine-elicited vnAbs demonstrate an *in vitro* neutralizing potency that is 2.76-fold higher against SARS-CoV-2 Wuhan compared to Delta variant. This *in vitro* relative potency, is lower than the 3.9 considered in the published model of protection^2^, which is likely due to either differences between *in vitro* neutralization levels and protection levels, or to the small sample size used in our testing that may not represent the full range of variation seen between individuals.

**Figure 2.**
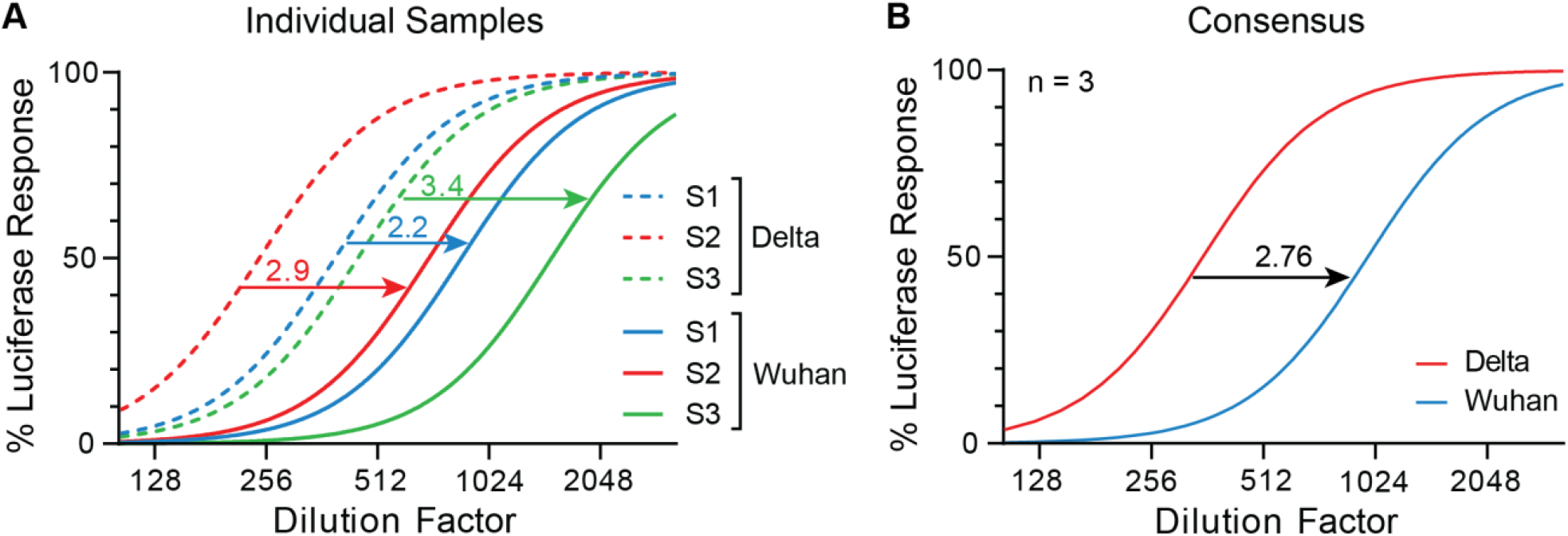
Comparison of vnAb neutralization of SARS-CoV-2 Wuhan and Delta. Three serum samples were tested side-by-side in modified IMMUNO-COV assays that specifically measure vnAb IC50 titers against SARS-CoV-2 Wuhan or Delta. Samples were tested in dilution series and the relative luciferase response curves (normalized to pooled seronegative serum) were used for regression analyses. Arrows indicated the fold-change in potency from SARS-CoV-2 Delta to Wuhan. A) Individual response curves for each serum sample. B) Consensus luciferase response curves.

Using internally collected historical data from IMMUNO-COV, we estimated the IMMUNO-COV vnAb titer needed for 50% protection against mild infection to be 67 IU/mL for SARS-CoV-2 Wuhan and 269 IU/mL for SARS-CoV-2 Delta (Figure 3A & C). Furthermore, we estimated the IMMUNO-COV vnAb titer needed for 50% protection against severe disease to be 10 IU/mL for Wuhan and 39 IU/mL for Delta (Figure 3B & D). The IMMUNO-COV limit of detection is 22 IU/mL. Thus, any detectable level of vnAbs likely confers some level of protection against severe disease, particularly in healthy individuals where cell-mediated adaptive immunity also helps reduce sickness from breakthrough infections. Of note, protection levels represent the degree that an individual’s baseline risk is reduced and do not account for underlying risk factors that an individual may already have from comorbidities.

**Figure 3.**
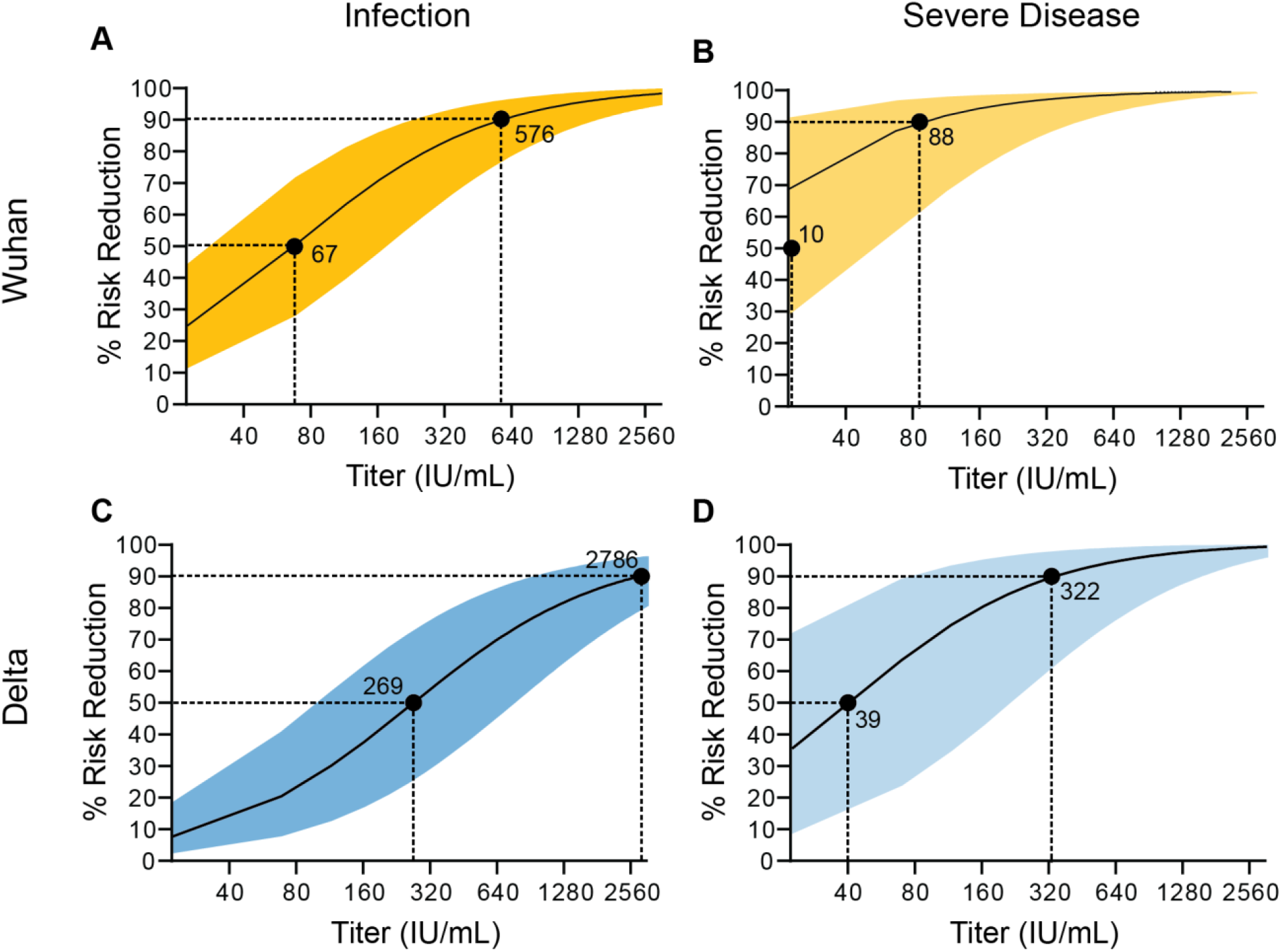
Models of protection. IMMUNO-COV historical titer data from convalescent individuals were applied to published models of protections from SARS-CoV-2 Wuhan (WT; ancestral) and Delta (B.1.617.2) variant. Protection is represented as percent risk reduction relative to vnAb titer as measured using the IMMUNO-COV assay. Models are shown for protection from SARS-CoV-2 Wuhan (A and B) and Delta variant (C and D), with regards to mild infection (A and C) or severe disease (B and D). The vnAb titers corresponding to 50% and 90% protection for each model are indicated.

### Longitudinal dynamics of vnAbs following vaccination

The longitudinal distribution of vnAb titers in participants followed a predicted trend, with individual titers initially rising post primary vaccination or vaccine boost and waning rapidly thereafter (Figure 4). Excluding titers boosted by COVID-19 breakthrough infection or booster vaccination, 29/52 (56%) participants maintained vnAb levels greater than 200 IU/mL out to six months post primary vaccination (five months post second vaccine dose). Based on our models, these titer levels provide strong (> 75%) protection against SARS-CoV-2 Wuhan infection, but less than 50% protection against infection by SARS-CoV-2 Delta. In the remaining 23/52 (44%) participants, vnAb titers dropped below 200 IU/mL between five to nine months post primary vaccination.

**Figure 4.**
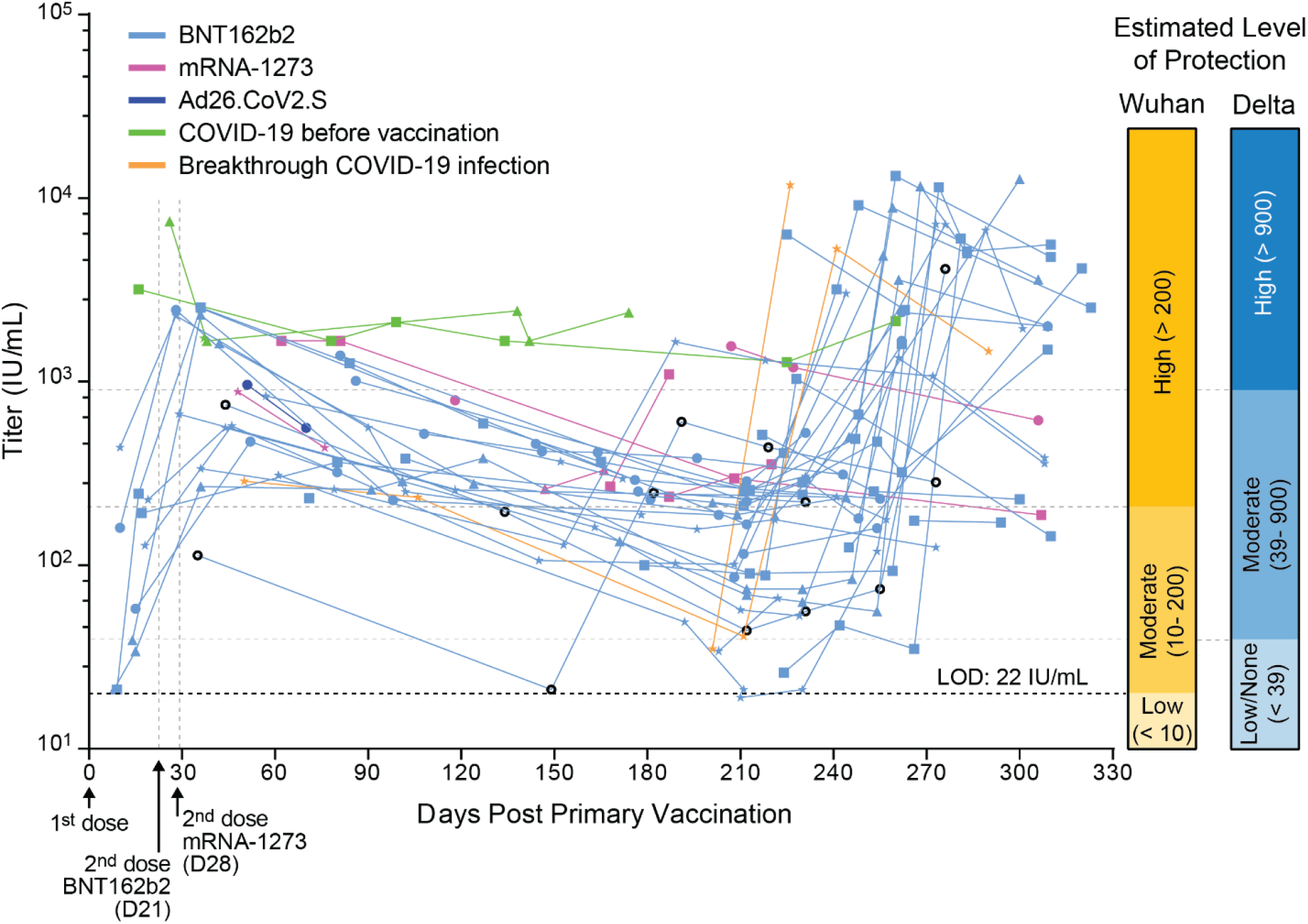
Longitudinal dynamics of vnAbs following primary, secondary and booster vaccinations. Longitudinal dynamics of vnAb titers in 56 participants are plotted over 330 days post primary vaccination (D0). Recommended time for secondary vaccination is indicated for BNT162b2 (D21) and mRNA-1273 (D28). Estimated levels of protection against SARS-CoV-2 Wuhan and Delta variant are indicated. Participants were vaccinated with BNT162b2 (light blue lines), mRNA-1273 (purple lines) or Ad26.CoV.S (dark blue lines). Four BNT162b2 vaccinees tested positive for COVID-19 prior to vaccination (green lines) and two participants had titers boosted by SARS-CoV-2 infection during the Delta surge (orange lines). Data is represented relative to participant age: 20-30 years old (circle); 31-40 years old (square); 41-50 years old (triangle); 51-60 years old (star); and above 61 years (torus). The IMMUNO-COV assay has a limit of detection of 22 IU/mL.

Peak vnAb titers were significantly higher following booster vaccination compared to primary vaccination. Modest increases in vnAb titers not attributable to breakthrough infection or booster vaccination were observed in 14/52 (27%) participants, primarily during the first SARS-CoV-2 Delta surge. Thus, these increases may have resulted from exposure or asymptomatic infections that were not recognized by participants, though variability in sample handling, or run-to-run assay variability may have also contributed to modest titer increases.

Several studies have reported that SARS-CoV-2 vnAb titers in convalescent individuals^13–15^ and vaccinees^16,17^ show rapid early decay that slows over time. This trend is consistent with a burst of short-lived antibody secreting cells producing peak vnAb titers, which decay rapidly until reaching a slowly declining plateau maintained for months to years by long-lived memory B cells. In our study, vnAb titers displayed similar kinetics, with titers in over half (32/52; 62%) of the participants reaching apparent homeostasis as early as three months post primary vaccination (Figure 4). When the kinetics of vnAb decay were analyzed in 60-day intervals (Figure 5), the half-life of vnAb decay averaged 1.77 months from the first two months to four months following vaccination. Because only 16/56 (29%) participants had titers measured within 8-30 days post second vaccination, peak titers were likely underestimated for many participants, and therefore, the rate of decay from two to four months is likely also underestimated. From four to six months following vaccination, the half-life of vnAb decay was 2.17 months, which dropped to 1.60 months from six to eight months post vaccination. This reduction in mean half-life was attributed in large-part to the 62% of participants that reached vnAb homeostasis, as the half-life for individuals who had not yet reached homeostasis was much higher (2.43 months compared to 1.08 months for individuals reaching homeostasis). Interestingly, individuals that reached homeostasis within six months, were primarily those with higher peak titers following vaccination. Comparison of the rate of decay between the three vaccine platforms was not performed due to insufficient longitudinal collections from mRNA-1273 and Ad26.CoV.S vaccinees.

**Figure 5.**
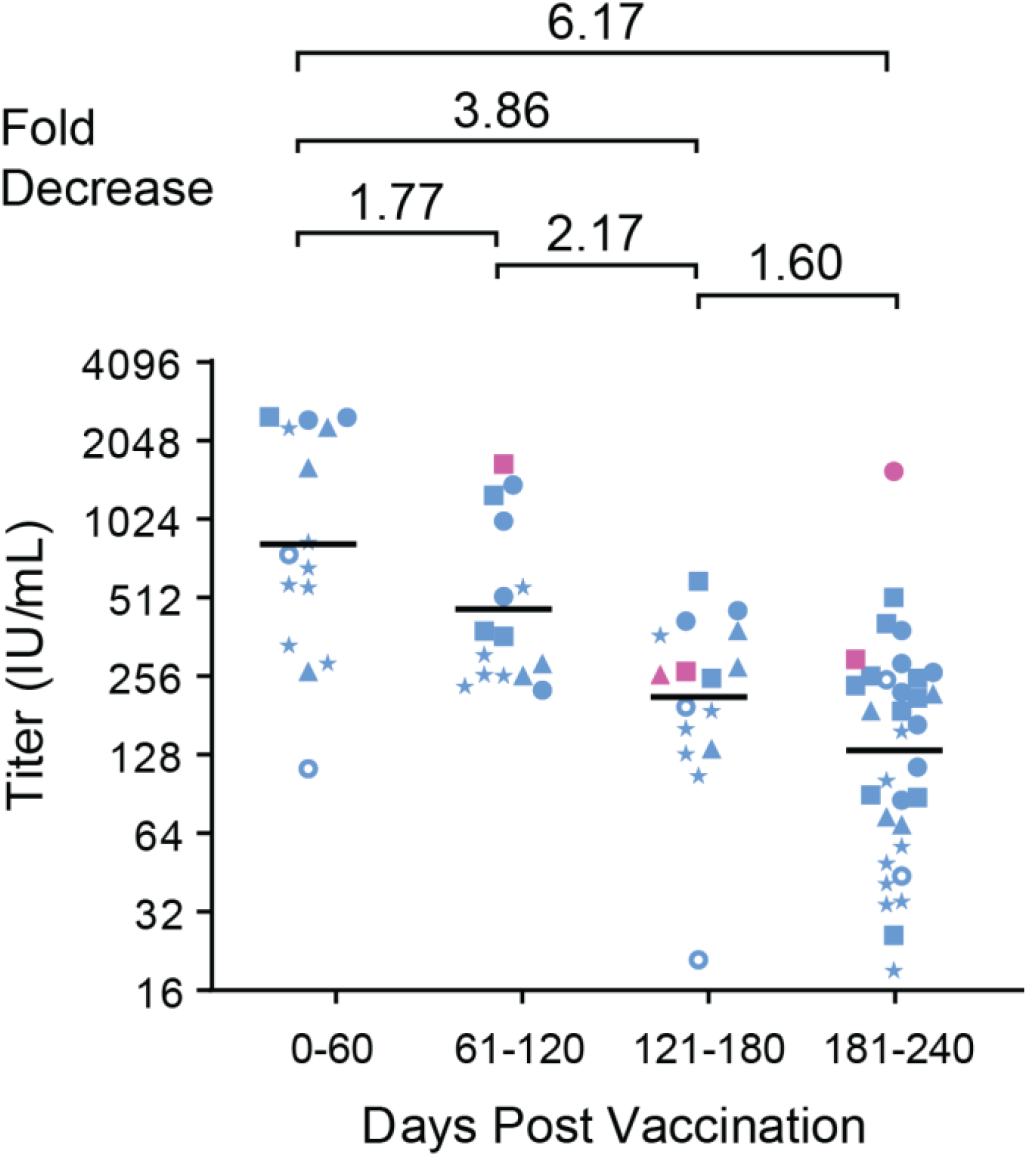
Longitudinal analysis of vnAb decay over 10 months. Kinetics of vnAb decay were plotted in 60-day intervals, and the fold decrease in geometric mean titer between each interval is shown. Participants convalescent from COVID-19 prior to vaccination were excluded from analysis, as were boosted titers following breakthrough infection or vaccine booster. Data is presented relative to participant age and vaccine received: BNT162b2 (blue); mRNA-1273 (purple); 20-30 years old (circle); 31-40 years old (square); 41-50 years old (triangle); 51-60 years old (star); over age 61 (torus).

### Correlation between Age and vnAb titer

A moderate negative correlation of statistical significance between vnAb levels and age was observed (Pearson’s coefficient, R −0.3552, P-value 0.0025) (Figure 6). When the correlation between age and vnAb levels was calculated per 60-day intervals, the negative correlation was more dramatic at later time points (Table 3), suggesting that the rate of vnAb decay increases with age. Consistent with this observation, the mean age of participants that maintained vnAb titers above 200 IU/mL over the course of the study (mean age 36 yrs, range 22-64 yrs) was lower than the mean age of participants whose vnAb dropped below 200 IU/mL (mean 44 yrs, range 24-64).

**Table 3:** vnAb titers relative to age at 2-month intervals following second vaccine dose

| Days post vaccination | D0-60 | D61-120 | D121-180 | D181-240 |
| --- | --- | --- | --- | --- |
| **Pearson r** | -0.3706 | -0.4356 | -0.6235 | -0.5235 |
| **95% confidence interval** | -0.7059 to -0.1006 | -0.7428 to -0.02323 | -0.8549 to -0.1849 | -0.7270 to -0.2354 |
| **R squared** | 0.1373 | 0.1897 | 0.3887 | 0.274 |
| **P value (two-tailed)** | 0.1183 | 0.0623 | 0.0099 | 0.0011 |
| **Total n** | 19 | 19 | 16 | 36 |

**Figure 6.**
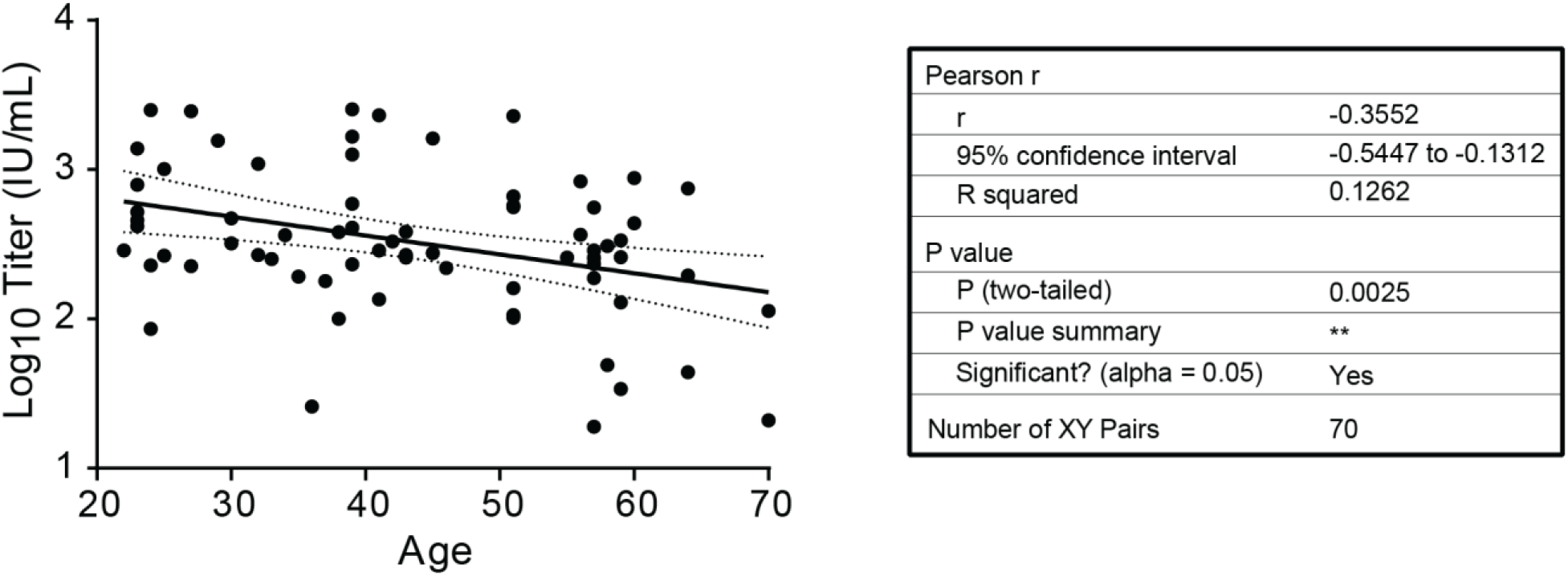
Correlation between age and titer. Correlation between age and log10-transformed vnAb titers was assessed. Pearson correlation and a simple linear regression are shown.

### Booster shots and secondary infections

Booster vaccinations were received by 37 (66%) participants. For 24 of the participants, vnAb titers were measured pre-boost and post-boost, and booster vaccination was associated with a mean 60.2-fold (range: 4.7 – 336.6) increase in vnAb titer (Figure 4 and Table 4). In 23/24 (96%) participants, boosted vnAb titers peaked above 900 IU/mL, corresponding to a predicted level of over 75% protection from infection and over 90% protection from severe disease caused by SARS-CoV-2 Delta. However, vnAb titers were transient and began to decay shortly after reaching peak levels (Figure 4). Two participants (3.6%), whose vnAb titers had dropped to 32 IU/mL and 27 IU/mL post primary vaccination, experienced breakthrough SARS-CoV-2 infections during the first Delta surge (Figure 4, orange lines). The breakthrough infections also provided an effective boost in vnAb titer that was comparable to the boost in titer observed following booster vaccination.

**Table 4:** Change in vnAb titers post booster vaccinations*

| Participant | Titer before boost (IU/mL) | Peak recorded titer post boost (IU/mL) | Days post boost to peak titer | Fold Change |
| --- | --- | --- | --- | --- |
| 1 | 21 | 605 | 14 | 28.8 |
| 2 | 35 | 6014 | 14 | 171.8 |
| 3 | 35 | 11781 | 17 | 336.6 |
| 4 | 53 | 1347 | 15 | 25.4 |
| 5 | 56 | 4111 | 23 | 73.4 |
| 6 | 56 | 11562 | 14 | 206.5 |
| 7 | 84 | 3588 | 15 | 42.7 |
| 8 | 86 | 4850 | 17 | 56.4 |
| 9 | 88 | 1035 | 10 | 11.8 |
| 10 | 93 | 11469 | 14 | 123.3 |
| 11 | 101 | 3023 | 33 | 29.9 |
| 12 | 119 | 7199 | 15 | 60.5 |
| 13 | 125 | 4142 | 42 | 33.1 |
| 14 | 129 | 1645 | 14 | 12.8 |
| 15 | 167 | 2389 | 17 | 14.3 |
| 16 | 177 | 6674 | 15 | 37.7 |
| 17 | 180 | 1669 | 14 | 9.3 |
| 18 | 234 | 8871 | 14 | 37.9 |
| 19 | 242 | 1595 | 16 | 6.6 |
| 20 | 257 | 13186 | 13 | 51.3 |
| 21 | 305 | 7167 | 16 | 23.5 |
| 22 | 321 | 1492 | 12 | 4.7 |
| 23 | 408 | 9146 | 14 | 22.4 |
| 24 | 499 | 12660 | 19 | 25.4 |
| **Mean** | 161 | 5718 | 17 | 60.2 |
| **SD** | 124.7 | 4163.3 | 6.9 | 77.7 |
| **Max** | 499 | 13186 | 42 | 336.6 |
| **Min** | 21 | 605 | 10 | 4.7 |

\*Data from 24 participants who gave blood samples pre- and post-vaccine boost.

### Effect of knowing vnAb titers on individual behaviors

To evaluate how real-time knowledge of vnAb titers impacted individual’s behavior and personal health decisions, a survey was sent to study participants at the end of 2021. A total of 44 (79%) participants returned results for the survey. Most participants (82%) considered knowing their individual vnAb titer as a positive experience (Figure 7A). The two individuals who had a negative impression of knowing their vnAb titer, indicated the information was useful, but disappointing due to their waning titer over time. The majority of participants (66%) also indicated that knowing their vnAb titer had a moderate to significant impact on their social behaviors (Figure 7B), including their practice of mask wearing in high-risk areas or activities and their participation in travel or family/social gatherings. Importantly, 66% of participants indicated that knowing their vnAb titer impacted their decision on when or if to get a booster vaccine (Figure 7C). Approximately, 23% of participants reported that they would like to repeat the test every six months, whereas the majority (75%), preferred a more frequent testing schedule (Figure 7D).

**Figure 7.**
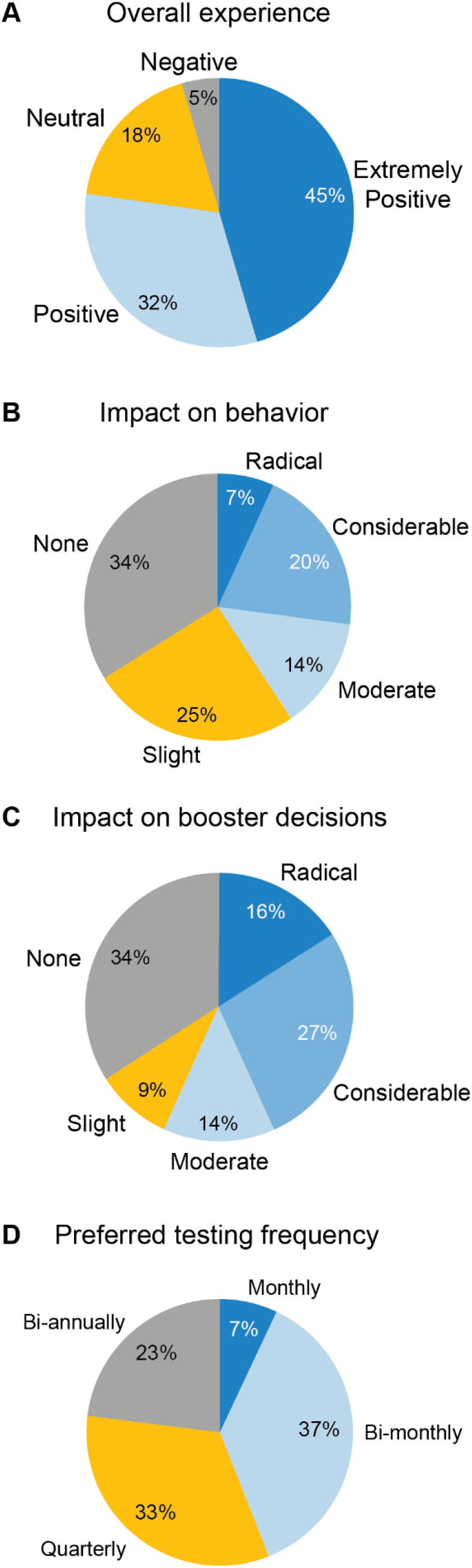
Knowing vnAb titers positively impacts personal wellness decision making. Results from an end of study survey of participants regarding how knowing vnAb titers impacted their behaviors and decisions (n = 44 responding). A) Participant overall view on knowing their vnAb titers. B) Impact knowing vnAb titer had on participant behaviors. C) Impact knowing vnAb titer had on participant decisions for timing of booster vaccination. D) Frequency of vnAb testing preferred by participants.

## DISCUSSION

Starting from January 1, 2021, employees from two companies were offered free access to IMMUNO-COV testing for determination of their vnAb titers against SARS-CoV-2. The goal of offering free testing was to provide a unique opportunity for participants to monitor their response to vaccination or natural infection, track their vnAb titers, and estimate their level of protection from future infection or disease in real time. The results of this study underscore the variability in vnAb titers and rates of decay among vaccinated individuals and highlight the need for frequent vnAb testing in personalized health assessments and behavioral decision making. Indeed, most study participants considered the knowledge of personal antibody titers to be helpful and used their vnAb titers to guide decisions regarding social gatherings and timing of booster vaccination.

Throughout the study year (2021), there were numerous local and federal recommendations and/or mandates addressing matters such as masking, social distancing, quarantining and vaccination. Individuals were specifically discouraged from using antibody testing to determine their own immune status and from making personalized decisions on that basis. Notably, the FDA issued a strongly worded recommendation in May 2021, discouraging the use of antibody testing as a means to determine the adequacy of a person’s immune response to vaccination and as a measure of their protections from reinfection^18^. This recommendation still stands at the time of writing this manuscript (February, 2022), despite publication of several pivotal peer-reviewed manuscripts and meta-analyses confirming vnAb titers provide an excellent correlate of protection from SARS-CoV-2 infection^1–4^. Indeed, there is now little argument in the scientific community that SARS-CoV-2 specific vnAbs prevent and/or mitigate SARS-CoV-2 infection and provide an excellent lab correlate of protection from COVID-19.

Unfortunately, most clinically approved antibody tests lack the ability to measure vnAbs, the antibody class that has been shown to correlate with protection. Instead, they detect – often qualitatively – antibodies capable of binding to the SARS-CoV-2 spike glycoprotein or nucleocapsid^19–21^. Because many binding antibodies are incapable of preventing virus infection, the information provided by such tests can be informative, but does not correlate as strongly to levels of protection as vnAb titers. IMMUNO-COV is a functional assay that uses a surrogate virus to specifically quantitate the titer of antibodies capable of preventing SARS-CoV-2 infection of cells. Thus, IMMUNO-COV titers provide a much better correlate of protection than other commercial antibody tests.

Studies have used a variety of assays to measure vnAb titers, including plaque reduction neutralization test (PRNT) with clinical isolates of SARS-CoV-2, pseudo-typed lentiviral assays, and pseudovirus VSV assays. These assays have used different approaches to quantitate vnAb levels. To facilitate cross comparisons from studies, publications have increasingly reported vnAb titers in IU/mL, based on calibration of the assays to the WHO International Standard^10^. On this basis, we calibrated IMMUNO-COV titers (VNT) to the WHO International Standard (Figure 1 and Table 2) and report here, our vnAb titer data in IU/mL. Due to availability of the WHO Reference Panel, calibration activities were not performed until fall 2021, and therefore, participants primarily received vnAb titer results in VNT units for this study.

While vnAb titers provide a good correlate of protection from early infection, we received informal feedback throughout our study that some participants had difficulty interpreting their vnAb titer results. Similar feedback was also received from some patients who ordered IMMUNO-COV testing through their physicians. To address this concern and provide individuals with a resource for understanding how their vnAb titer relates to estimated protection, we applied historical IMMUNO-COV titers from convalescent individuals to models of protection reported by Khoury *et al*.^1^. IMMUNO-COV specifically measures vnAb titers against ancestral SARS-CoV-2 Wuhan, and therefore, the initial modeling was done to correlate vnAb titer to protection from Wuhan (Figure 3). Because vnAbs demonstrate varying degrees of cross protection from SAR-CoV-2 viral variants, models of protection must be adjusted for each viral variant^2^. We also modeled IMMUNO-COV vnAb titer correlations with protections from SARS-CoV-2 Delta, which became the dominant strain within the U.S.A. during the course of this study. Models for SARS-CoV-2 Delta were based on a published conversion factor of 3.9^2^, which was similar to the relative potency factor of 2.76 that we calculated from a small sample set run side-by-side in IMMUNO-COV and IMMUNO-COV (Delta) assays (Figure 2). Thus, for SARS-CoV-2 Delta, higher vnAb titers are required to provide protection when compared to Wuhan. While not directly modeled here, available scientific evidence^22–26^ indicates that similar models of protection from the now dominant Omicron variant will be shifted even higher.

In the current study, all participants developed neutralizing antibodies post vaccination after receiving BNT162b2, mRNA-1273, or Ad26.CoV.S vaccines (100% seroconversion rate). Over half (62%) of participants appeared to reach a state of vnAb homeostasis over six months post second vaccine dose (Figure 4). Additionally, in 56% of participants, vnAb titers remained above 200 IU/mL, representing an estimated 75% protection against infection from SARS-CoV-2 Wuhan (Figure 3). Consistent with previous reports,^17,27,28^ we observed a negative correlation between vnAb titers and age (Figure 6). Older participants on average were less likely to maintain titers above 200 IU/mL and demonstrated more rapid rates of vnAb decay. Most participants who received booster vaccines were previously vaccinated with BNT162b2 and received a homologous booster vaccine (Table 1). All individuals experienced an increase in vnAb titers after booster vaccination (Figure 4). Interestingly, individuals with pre-boost titers below 200 IU/mL demonstrated slightly larger fold increases in vnAb titers following booster vaccination but lower on average post-boost peak titers relative to individuals with pre-boost titers above 200 IU/mL (Table 4).

Through our study, participants had a unique opportunity to use their real-time vnAb titers to estimate their level of protection from future infection and use that information to aid in personal decision making during the pandemic. Survey results indicated that most participants considered knowledge of vnAb titers helpful, with respondents citing peace of mind, impact on social distancing decisions – especially in relation to large social gatherings, and impact on decisions regarding the timing of booster vaccinations. With the most recent SARS-CoV-2 Omicron surge, reports of less severe disease, and growing pandemic weariness, a higher percentage of healthy individuals within the U.S.A. are less concerned about future infection. Yet, for individuals with high risk of severe disease – such as those with comorbid conditions or weakened immune systems – and their families, making medical and behavioral decisions to reduce possible SARS-CoV-2 infection remains a priority. For this group of people especially, tests like IMMUNO-COV that can provide individuals with an estimate of their level of protection have great utility. Given the dramatically reduced susceptibility of SARS-CoV-2 Omicron to vaccine-induced vnAbs, we are currently validating a new assay, IMMUNO-CRON™. IMMUNO-CRON specifically quantifies vnAb titer in response to SARS-CoV-2 Omicron, and therefore, can be used to better estimate protection against the currently dominant variant. Once clinically validated, IMMUNO-CRON will be available through physician order, to continue providing individuals with vnAb information that can aid in their personal health decisions.

## Data Availability

All data produced in the present work are contained in the manuscript.

## Acknowledgements

We would like to thank the employees who participated in this testing program and gave consent to use their data as part of this retrospective study.

## Conflicts of interests

Authors have a financial interest due to equity in Imanis Life Sciences (Mayo Clinic, SJR, KWP) and Vyriad (SJR, KWP, SN), or are employees of Imanis Life Sciences or Vyriad.

## Author Contributions

Designed and planned study and experiments: RV, TC, L. Suksanpaisan, LR, SN, PL, KWP, SJR; performed experiments and testing: CL, RN, MH, L. Schnebeck, AW, AD; analyzed data: RV, TC, L. Suksanpaisan, CL, RN, PL; wrote the manuscript: RV, PL, SJR

## Notes

### Funding Statement

This study was funded internally Vyriad, Inc. and Imanis Life Sciences.

### Author Declarations

The study protocol, informed consent forms to collect blood, and participant survey were reviewed and approved by Western Institutional Review Board.

